# SARS-CoV-2 Seroprevalence among Healthcare Workers

**DOI:** 10.1101/2021.10.02.21264468

**Authors:** Talia D. Wiggen, Bruno Bohn, Angela K. Ulrich, Steven D. Stovitz, Ali J. Strickland, Brianna M. Naumchik, Sara Walsh, Stephen Smith, Brett Baumgartner, Susan Kline, Stephanie Yendell, Craig Hedberg, Timothy J. Beebe, Ryan T. Demmer

## Abstract

**Background:** Monitoring COVID-19 infection risk among health care workers (HCWs) is a public health priority. We examined the seroprevalence of SARS-CoV-2 among HCWs following the fall infection surge in Minnesota, and before and after COVID-19 vaccination. Additionally, we assessed demographic and occupational risk factors for SARS-CoV-2 infection.

**Methods:** We conducted two rounds of seroprevalence testing among a cohort of HCWs: samples in round 1 were collected from 11/22/20 - 02/21/21 and in round 2 from 12/18/20 - 02/15/21. Demographic and occupational exposures assessed with logistic regression were age, sex, healthcare role and setting, and number of children in the household. The primary outcome was SARS-CoV-2 IgG seropositivity. A secondary outcome, SARS-CoV-2 infection, included both seropositivity and self-reported SARS-CoV-2 test positivity.

**Results:** In total, 459 HCWs were tested. 43/454 (9.47%) had a seropositive sample 1 and 75/423 (17.7%) had a seropositive sample 2. By time of sample 2 collection, 54% of participants had received at least one vaccine dose and seroprevalence was 13% among unvaccinated individuals. Relative to physicians, the odds of SARS-CoV-2 infection in other roles were increased (Nurse Practitioner: OR[95%CI] 1.93[0.57,6.53], Physician’s Assistant: 1.69[0.38,7.52], Nurse: 2.33[0.94,5.78], Paramedic/EMTs: 3.86[0.78,19.0], other: 1.68[0.58,4.85]). The workplace setting was associated with SARS-CoV-2 infection (p=0.04). SARS-CoV-2 seroprevalence among HCWs reporting duties in the ICU vs. those working in an ambulatory clinic was elevated: OR[95%CI] 2.17[1.01,4.68].

**Conclusions:** SARS-CoV-2 seroprevalence in HCW increased during our study period which was consistent with community infection rates. HCW role and setting — particularly working in the ICU — is associated with higher risk for SARS-CoV-2 infection.

## INTRODUCTION

The Coronavirus-2019 (COVID-19) pandemic, caused by severe acute respiratory syndrome coronavirus 2 (SARS-CoV-2) continues to pose a risk to healthcare workers (HCWs), particularly in global settings where vaccinations are not widely available. HCWs are vital in our response to COVID-19. Of COVID-19 case reports sent to the CDC early in the pandemic (February – April 2020) that included data on HCW status, 19% were identified as HCWs.^1^ Due to occupational exposure to infectious patients and materials, HCWs have the potential for increased exposure to SARS-CoV-2^2–6^ and prior studies indicate that HCWs are at increased risk of infection with SARS-CoV-2.^1,3–6^ Furthermore, occupational factors such as role, setting, and PPE availability have been shown to be associated with seroprevalence in HCWs.^2–4,7,8^ However, it remains unsettled whether these occupational features translate into higher risk of infection among HCWs relative to the risk of community transmission, and if so, which HCWs are at highest risk. Other studies have observed that HCWs have lower or equal infection risk to that of the community and report that hospital work was not associated with infection.^7,9^ This could possibly be explained by infection prevention strategies.^7,10^

A limitation of prior studies is the use of PCR testing, which only captures current infection. Testing for antibodies against SARS-CoV-2 can detect previous infection. Seroprevalence estimates in HCWs have been reported in multiple countries with a wide range of seropositivity levels that differ by region and stage of the pandemic: US, 1.3-27% between March-June 2020 ^2,5,7–9^; Spain, 9.3% between March-April 2020 ^11^; UK, 10.8-24.4% between April-July 2020^4,12^; Belgium, 6.4% in April 2020^13^; Germany, 1.6% between March-April 2020 ^10^; China, 3.8% between March-April 2020.^14^ The variation in results, highlights the importance of ongoing surveillance in a variety of settings to better identify risk dynamics among HCWs. Limited seroprevalence data are available from HCWs following the infection surge that occurred in the U.S. during the fall of 2020.

Additionally, the impact of vaccination on serosurveys among HCWs following the emergency use authorization issued on December 11, 2020^15^ in the U.S. has not been reported. To our knowledge, there are no population-level data to inform how well vaccination will be captured by standard IgG assays nor are there data on timing of seroconversion in real-world setting.

We conducted a cohort study to examine the seroprevalence of anti-SARS-CoV-2 IgG among health care workers in the Minneapolis area between November 2020 – February 2021. Additionally, we assessed demographic and occupational risk factors for SARS-CoV-2 infection among HCWs and evaluate seroprevalence before and after COVID-19 vaccination among HCWs.

## METHODS

### Study design and population

The cohort included HCWs from a cross-sectional convenience sample of individuals working in healthcare facilities located in the Minneapolis/St. Paul, MN metropolitan area. Participants were previously recruited and enrolled as part of an ongoing SARS-CoV-2 RT-PCR cohort study.^16^ Briefly, participants were identified via social media advertisements and enrolled from April 20^th^–June, 24^th^, 2020. Eligibility criteria were: i) employed or volunteering in a healthcare facility; ii) free of fever, chills, anosmia, pharyngitis, recently developed persistent cough, nasal congestion suspected to be unrelated to season allergies; iii) age 18-80 years; iv) not pregnant. Participants who enrolled and consented to participate in the previous study were then recruited via an email sent on 11/17/2020 to participate in a seroprevalence study. Participants were invited to provide up to 2 capillary blood samples for serology testing. Participants’ first samples (“sample 1”) were collected between 11/22/20 - 02/21/21 and participants’ second samples (“sample 2”) were collected between 12/18/20 - 02/15/21 (Figure 1A). The study was approved by the University of Minnesota Institutional Review Board (#STUDY00009404). All participants provided informed consent. Of the 483 participants that provided informed consent, 459 (95%) were included in the final analysis (Figure 1B).

**Figure 1A.**
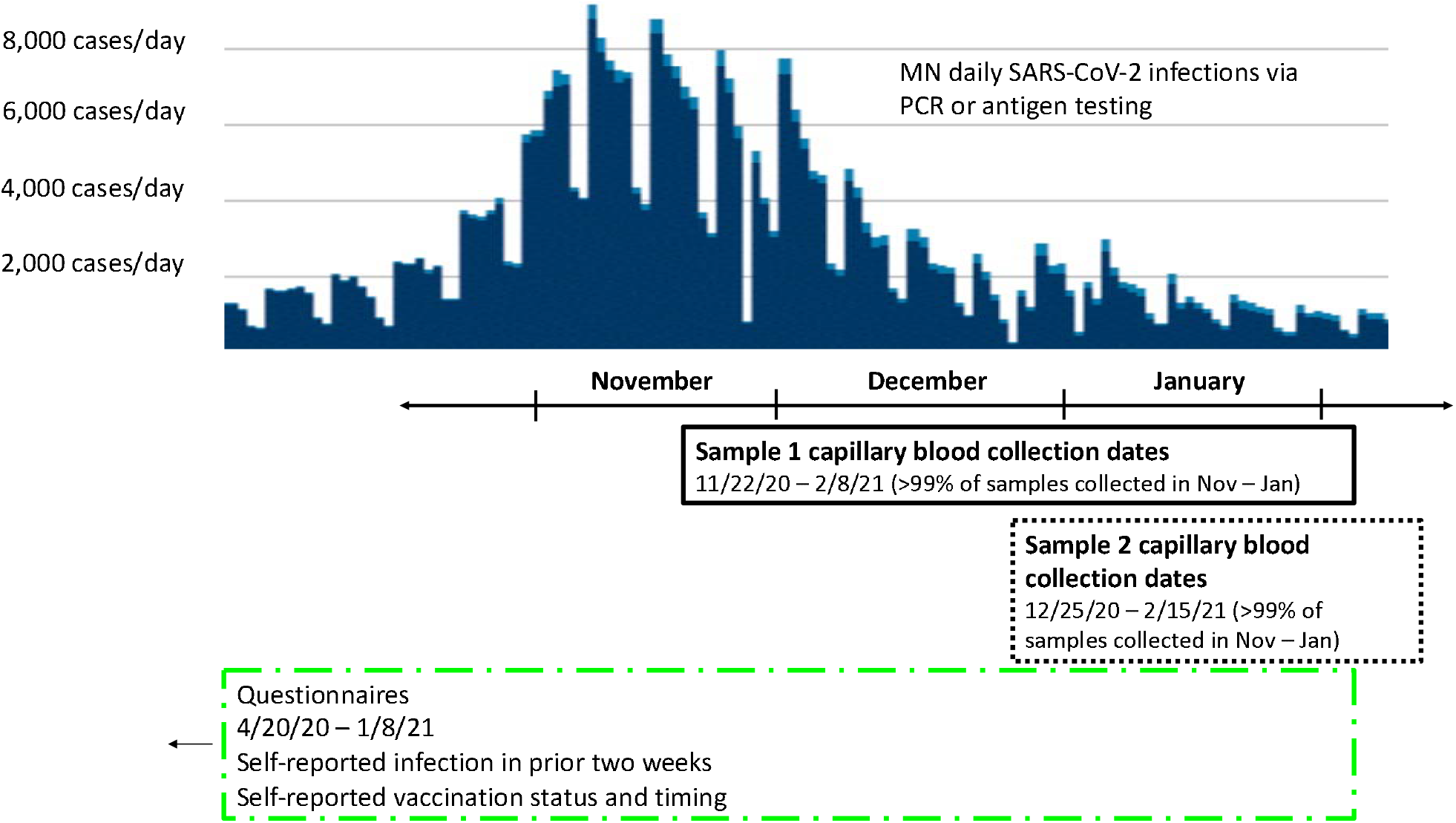
Study Design and Timing Overview.

**Figure 1B:**
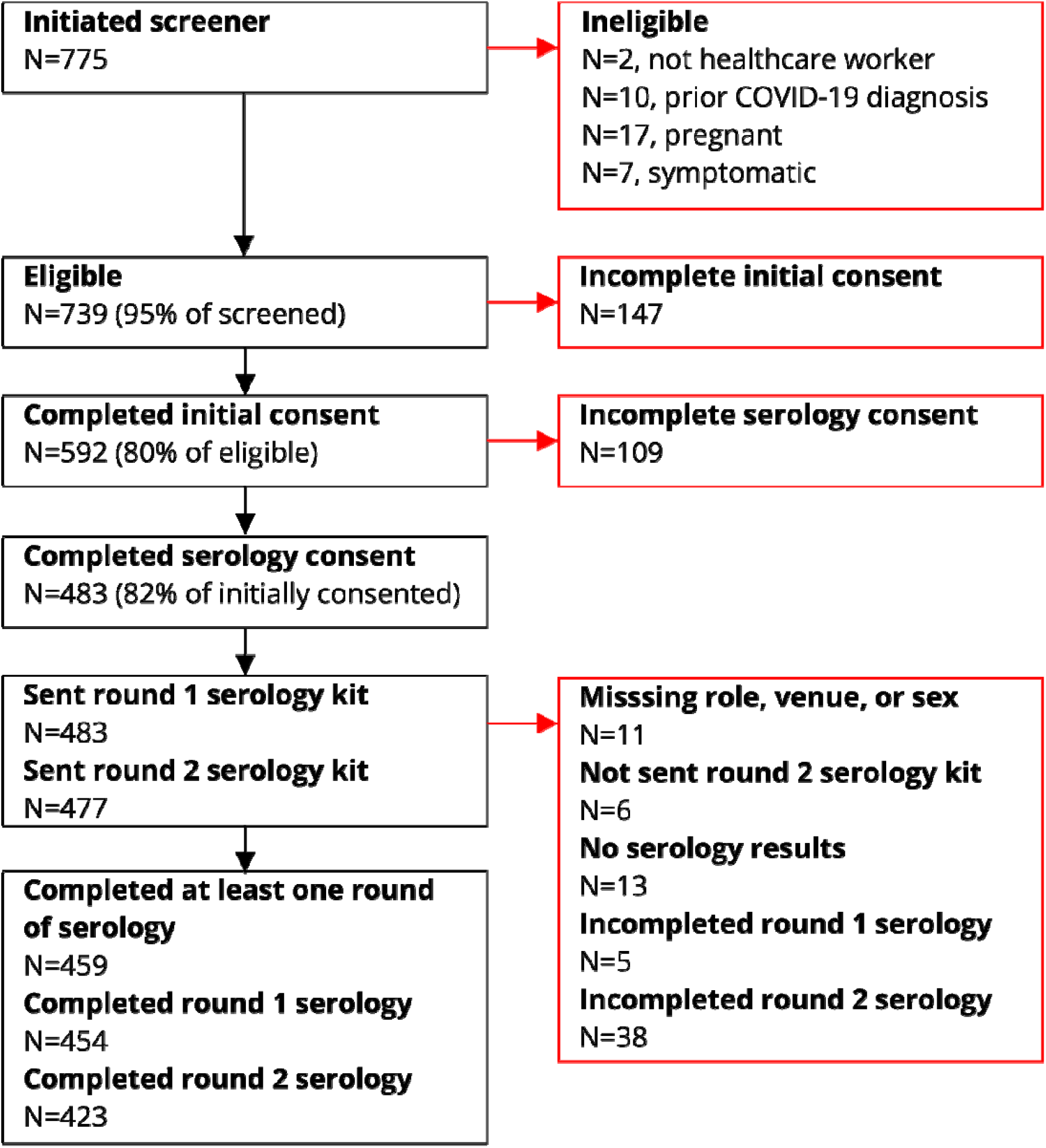
Participant flow diagram.

### Sample collection and antibody testing

Participants self-collected capillary blood using an at-home serology kit with a Mitra blood spot collection device (https://www.neoteryx.com/home-blood-blood-collection-kits-dried-capillary-blood). SARS-CoV-2 IgG antibody testing was performed at a central Quansys Bioscience laboratory using Quansys Biosciences’ SARS-CoV-2 Human IgG (4-Plex) Q-Plex multiplex assay, an enzyme-linked immunosorbent assay (ELISA).^17^ The Quansys SARS-CoV-2 Human IgG (4-Plex) ELISA tests for IgG antibodies to either SARS-CoV-2 spike proteins, S1 and S2. Quansys assay validation study reports an estimated sensitivity of 97% and specificity of 100%.^17^

The primary outcome of interest was SARS-CoV-2 IgG seropositivity. A secondary outcome, SARS-CoV-2 infection, was a combined variable including serology test results described above and self-reported previous positive SARS-CoV-2 PCR or antigen test used for associational analysis.

### Study variables

Participants self-reported data on age, sex, race, healthcare role and setting, number of children in the household, and comorbidities. Participants self-reported COVID-19 vaccination status including number of doses received, and date of each vaccine dose via biweekly surveys. Participants were able to select more than one work setting; for associational analysis, participants were assigned to the setting with the hypothesized highest risk of SARS-CoV-2 infection. Work setting from hypothesized highest to lowest risk was categorized by intensive care unit, emergency department, other inpatient, ambulatory clinic, emergency transport vehicle, and other setting.

### Statistical analysis

All statistical tests were performed uswing SAS University. Descriptive data are summarized as counts and percentages To account for testing error, adjusted seroprevalence was calculated using the formula: 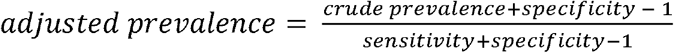 .^18^ To remove the influence of COVID-19 vaccination on seroprevalence trends, we present seroprevalence stratified by month of serology testing only among participants who were not vaccinated for COVID-19 at the time of serological testing (Figure 2B). Univariable logistic regression was used to examine the association between SARS-CoV-2 seropositivity and age, sex, work setting, work role, children in the household and vaccination status. Among a subgroup of participants that had not yet received the COVID-19 vaccine at the time of serology testing we also conducted logistic regression analyses to assess risk for infection (combining participant self-report of prior infection and seropositivity among unvaccinated individuals).

**Figure 2.**
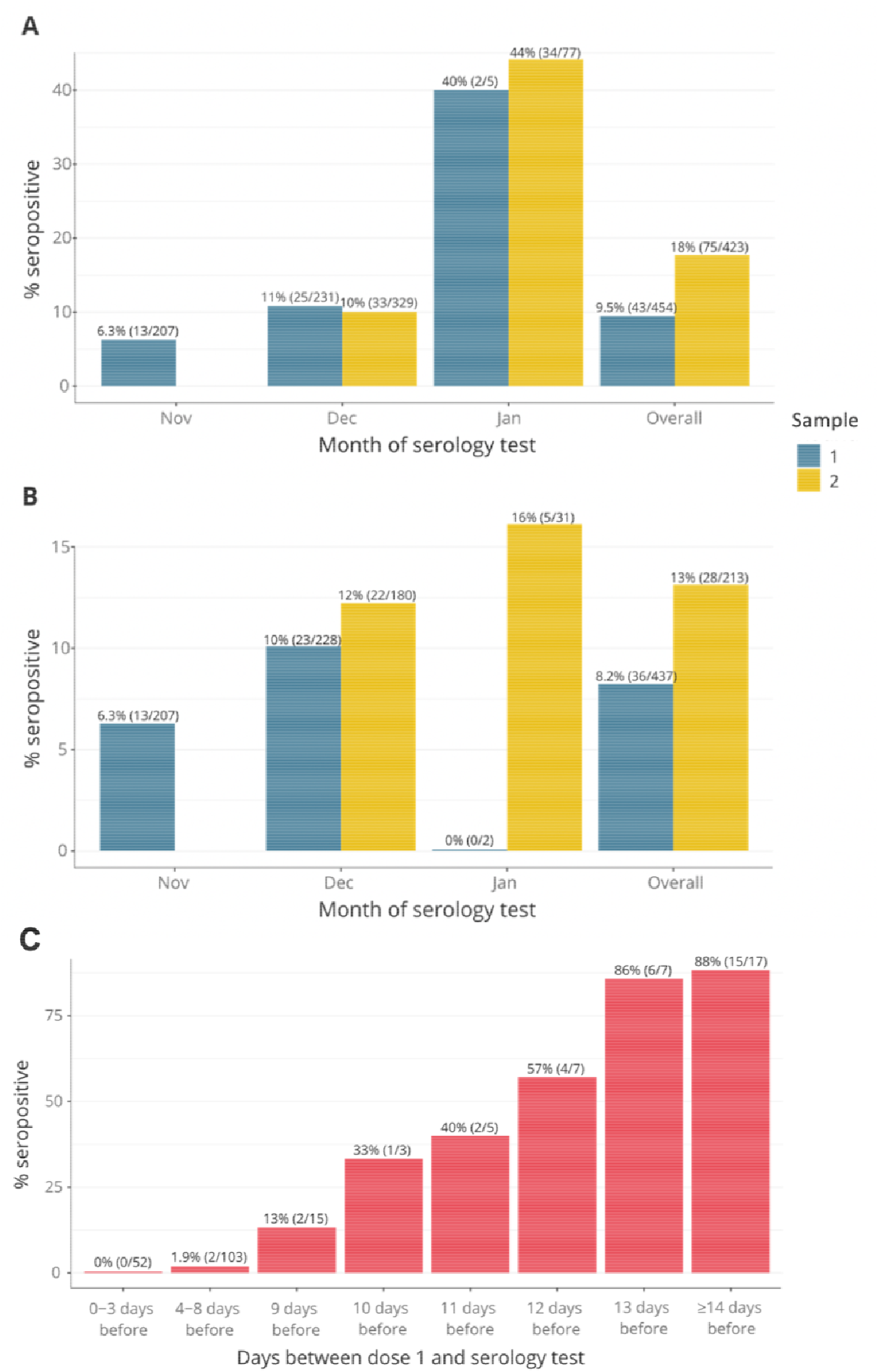
SARS-CoV-2 seroprevalence by month for sample 1 and sample 2 serology testing among **A)** all participants, **B)** among only those who were not vaccinated for COVID-19 prior to the day of serology testing, and **C)** according to time from vaccination among vaccinated individuals. %(N/total). In panel A, samples from February are included in the Overall group, but not presented separately due to low numbers. In sample 1 (N=1), 0% were seropositive, in sample 2 (N=5) 100% were seropositive. Samples with a missing date of collection are only included in the Overall group. In sample 1 (N=10), 30% were seropositive. In sample 2 (N=12), 25% were seropositive. In panel B, there are 2 samples from February or without a date of collection included in the overall results.

## RESULTS

### General population characteristics

Table 1 describes the study population characteristics and SARS-CoV-2 IgG seroprevalence. The majority of the participants were female (85%) and White (91%). Nurses (53%) and those working in a non-ICU inpatient setting (40%) made up the largest proportion of the participants. As of March 4^th^, 2021, 86% of the study population had received their first COVID-19 vaccine dose and 83% had received the second dose.

**Table 1.**
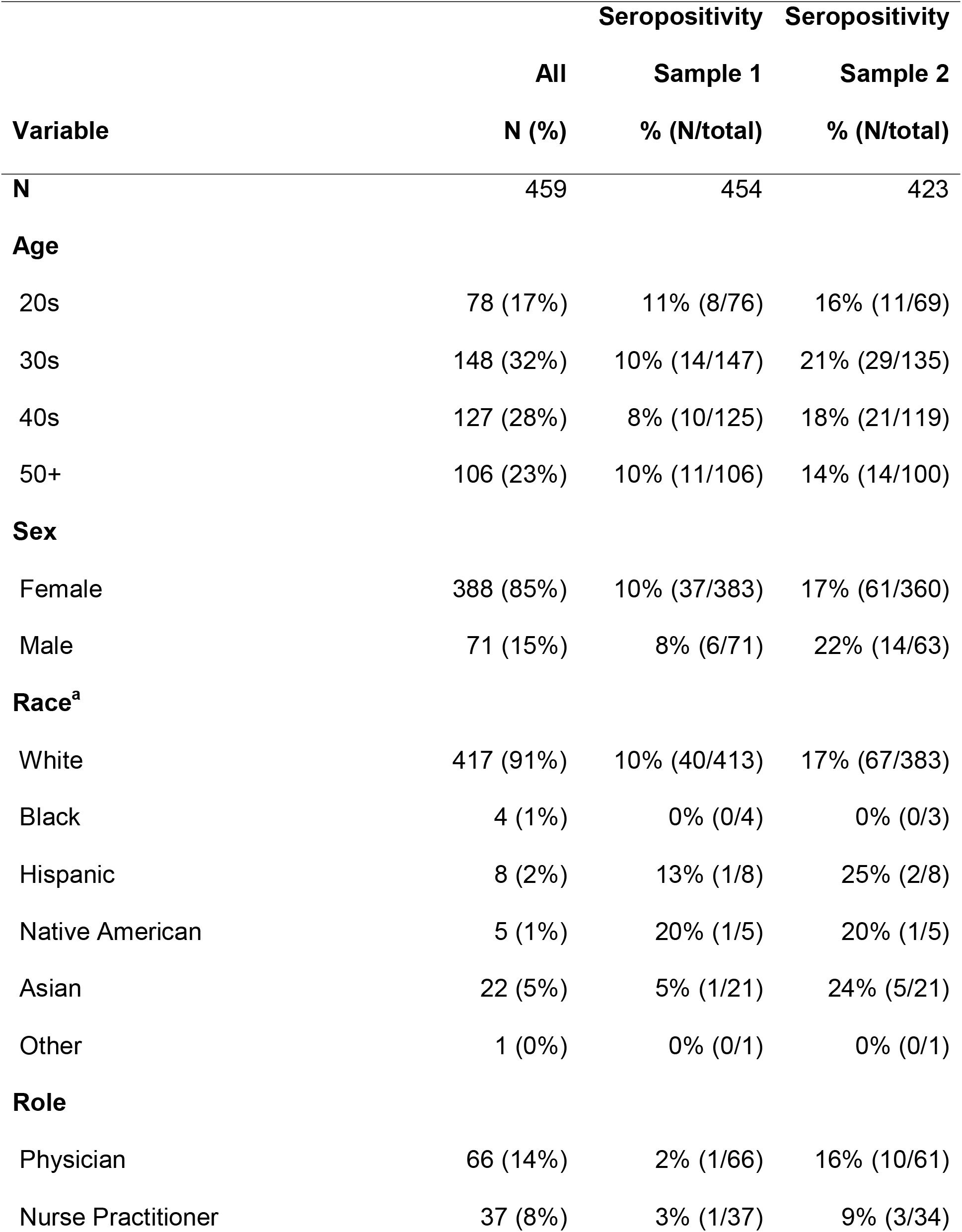
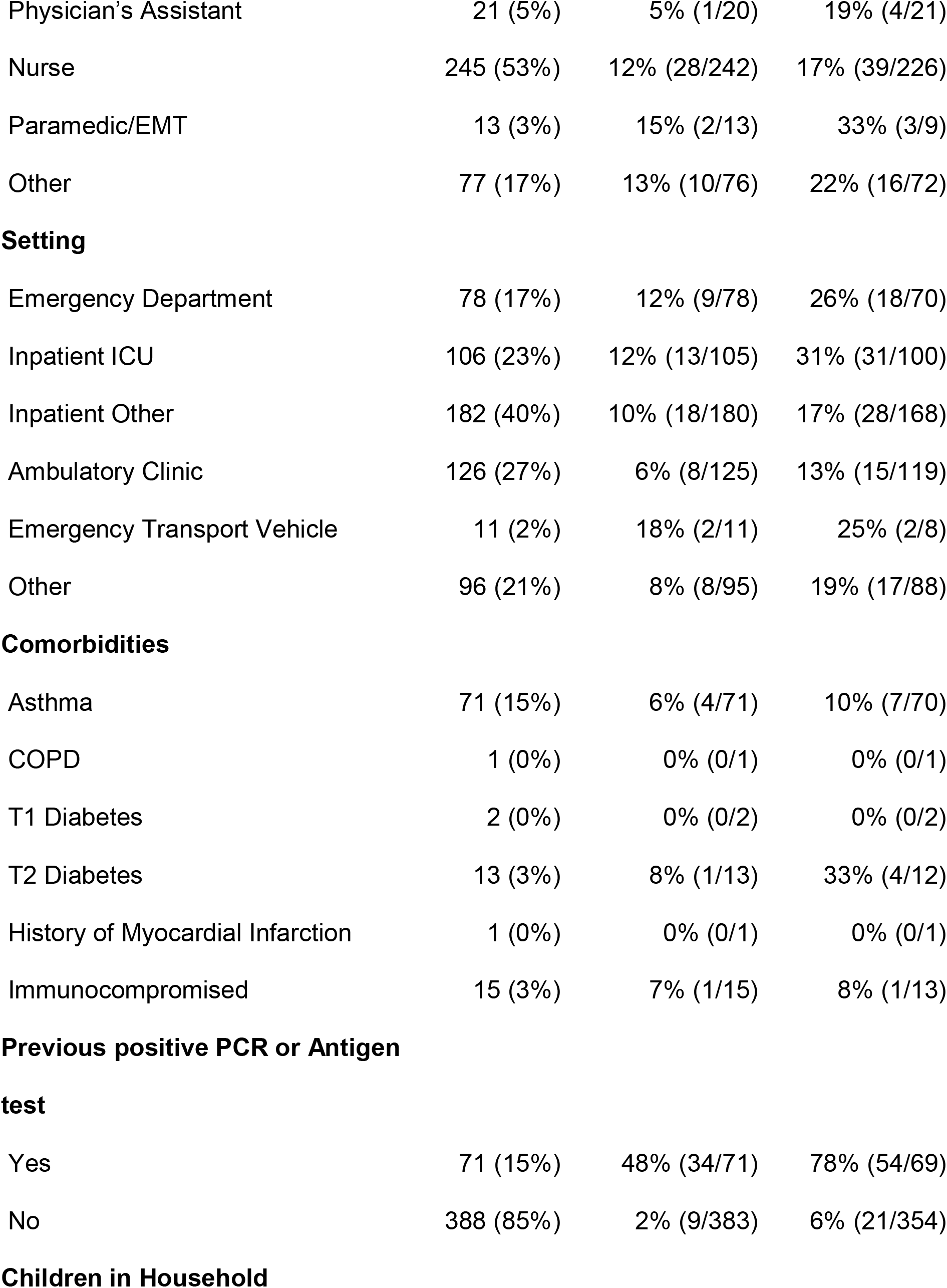
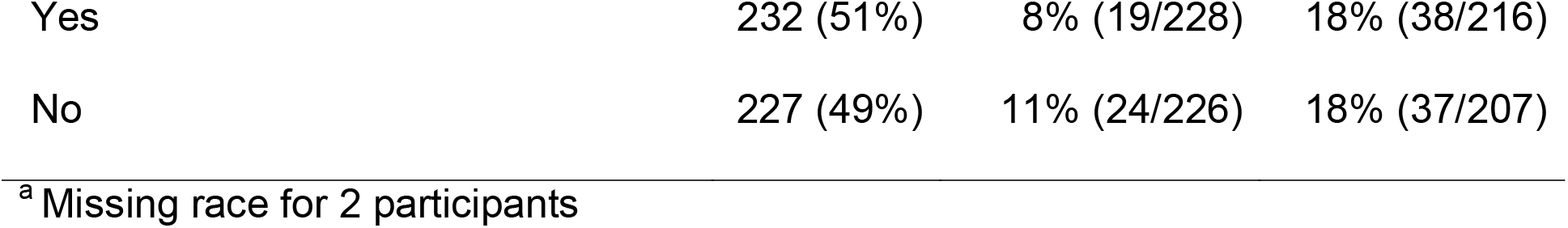
General participants characteristics and seroprevalence among each category.

### SARS-CoV-2 IgG seroprevalence

Among all participants, SARS-CoV-2 seropositivity was 9.5% (43/454, 95%CI: 6.9%-12.5%) for sample 1 and 17.7% (75/423, 95%CI: 14.2%-21.7%) for sample 2. The median time between sample 1 and sample 2 collection was 28 days with an interquartile range of 13. Testing error-adjusted SARS-CoV-2 IgG seroprevalences were 9.8% and 18.3%, respectively. Among unvaccinated participants, 8.2% (36/437, 95%CI: 5.8%-11.22%) and 13.1% (28/213, 95%CI: 8.9%-18.4%) were seropositive for samples 1 and 2, respectively. Seroprevalence stratified by month of serology testing is shown in Figure 2A. Seroprevalence increased from November to January among all participants (Figure 2A) and among those who were unvaccinated at the time of testing (Figure 2B).

### COVID-19 vaccination and SARS-CoV-2 IgG seroprevalence

Approximately half (209/412) of participants who reported their vaccination status were vaccinated prior to their second serology assay. Among those who reported receiving a first dose 0-3 days prior to testing, 0% of participants were seropositive while 88% were positive among those receiving their first vaccine dose at least 14 days prior (Figure 2C).

### Demographic and occupational risk factors associated with SARS-CoV-2 infection

To further explore the association between demographic and occupational factors with SARS-CoV-2 infection we assessed predictors of infection among individuals without prior vaccination. Role in the healthcare system and clinical setting were the two strongest predictors of prior infection (Table 2). Relative to physicians, the odds of SARS-CoV-2 infection were 1.93 (95%CI: 0.57, 6.53) in nurse practitioners, 1.69 (95%CI: 0.38, 7.52) in physician’s assistants, 2.33 (95%CI: 0.94, 5.78) in nurses, 3.86 (95%CI: 0.78, 19.0) in paramedic/EMTs, and 1.68 (95%CI: 0.58, 4.85) in other roles; pooled results comparing physicians to non-physicians were not statistically significant. Clinical setting was significantly associated with SARS-CoV-2 infection (p=0.03). The odds ratio for SARS-CoV-2 infection among those working in the ICU vs. the ambulatory setting was 2.17 (95%CI: 1.01, 4.68). After multivariable adjustment for all variables in Table 2, results were similar. The odds ratio for infection among HCWs with 3 or more kids in the household was OR:1.94 (95%CI: 0.81, 4.64) after multivariable adjustment as in Table 2,

**Table 2.**
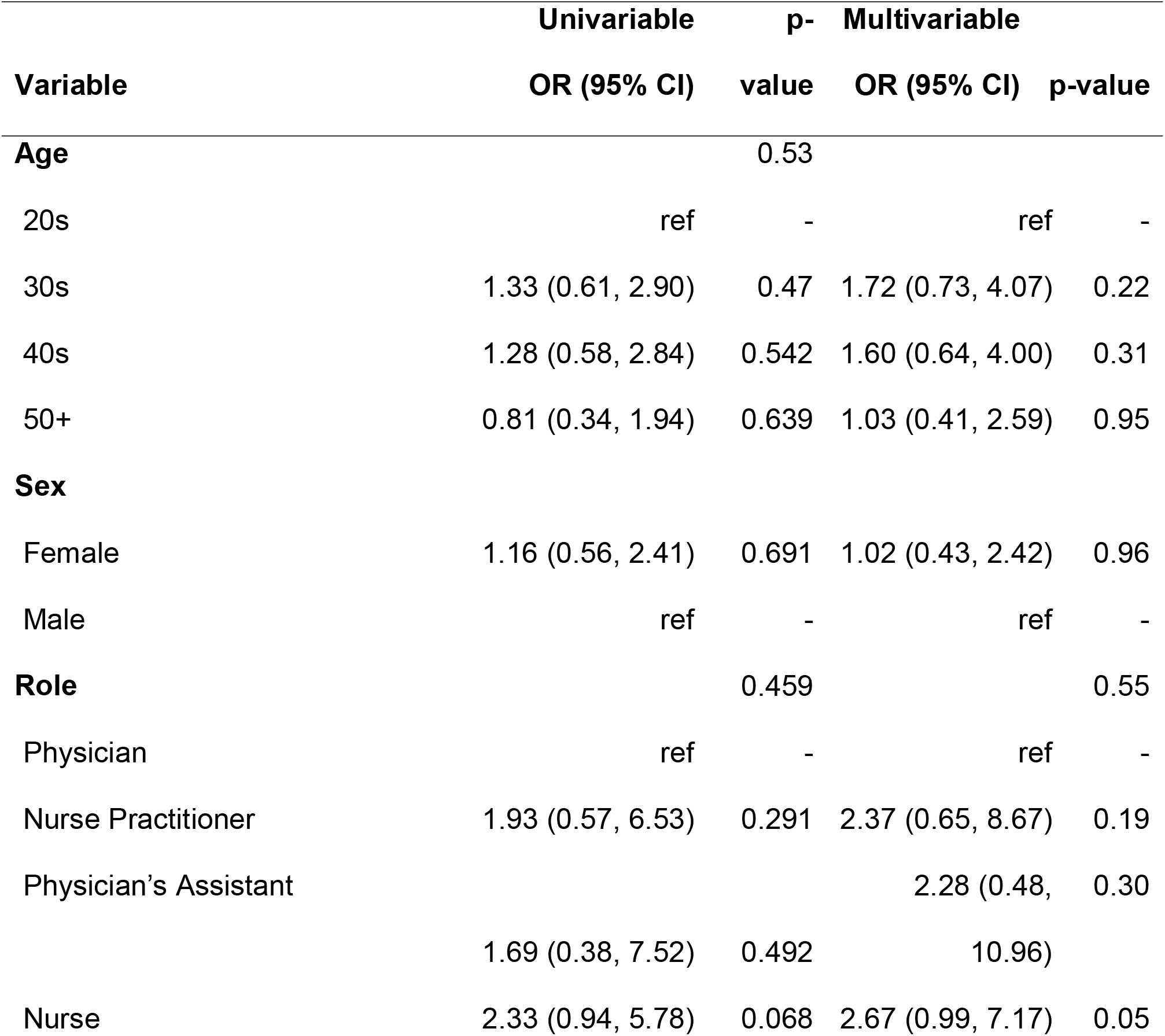
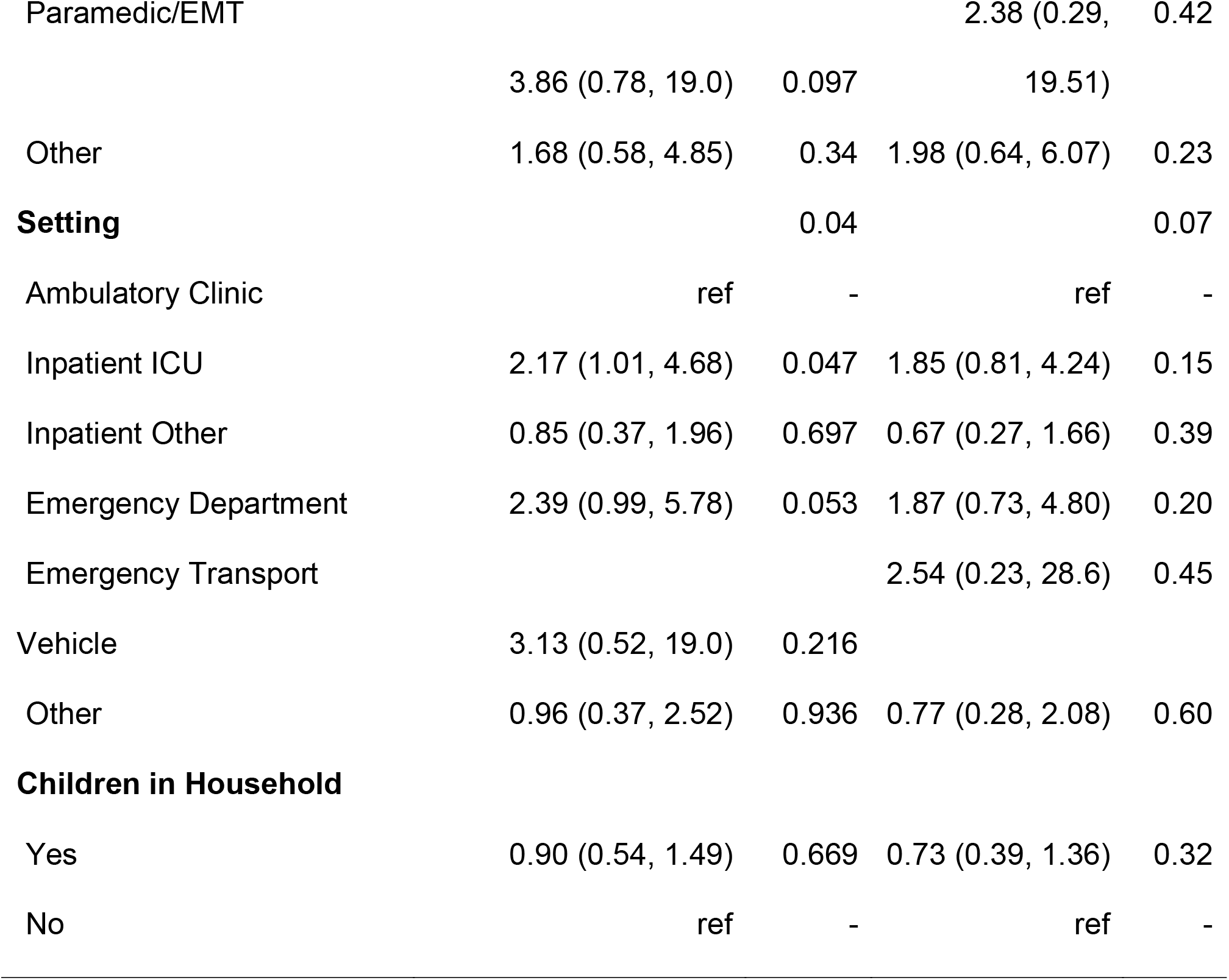
Association of SARS-CoV-2 infection with participant characteristics using logistic regression among subgroup of participants that completed a nasopharyngeal swab and were not vaccinated for COVID-19 at time of serology testing (N=402).

## DISCUSSION

In this study of HCWs, we found the SARS-CoV-2 IgG seroprevalence from sample 1 to be 9.5% and from sample 2 to be 17.7%. Seroprevalence increased ∼4% per month and these trends were independent of vaccination. Working in the ICU was associated with higher risk of prior infection and non-physicians realized an empirically higher risk of infection although this was not statistically significant.

In our convenience sample of healthcare workers, the seroprevalence of SARS-CoV-2 was similar to estimates in the general Minnesota population; Minnesota showed an approximately four-fold increase in PCR confirmed cases during the fall.^19^ Our absolute estimates are also similar to CDC results from Minnesota which estimated seroprevalence to be 11.4% (95%CI: 9.2%-13.8%) in November 2021 and 15.9% (95%CI: 13.3%-18.6%) in the first half of January 2021.^20^

While our data have no ability to know whether infections were community or occupationally derived, they are consistent with several^7,9, 24^ but not all^4^,^6^ prior studies also reporting similar infection rates between HCWs overall and the general community. Variation in findings could be due to several reasons including the stage of the pandemic at which the study was performed, variable community prevalence of SARS-CoV-2, differing infection control policies, and access to appropriate PPE. Additionally, understanding the makeup of the HCW sample is important as HCWs with direct patient care responsibilities, particularly for SARS-CoV-2 infected patients, clearly carry a higher risk for occupational exposure and infection compared to those in roles without direct patient contact. This is supported by our findings that work in the ICU was related to an increased risk of infection. The increased risk from working in the ICU is likely linked to increased exposure to COVID-19 patients or possibly exposure to aerosols generated by patient ventilation. We previously observed that 30% of participants working in ICUs reported a direct COVID-19 exposure within the past two weeks as compared to 6% of HCWs in ambulatory clinical settings^16^. Other cross-sectional serosurveillance studies have found no association between setting and seropositivity^21^. And some have reported that working in the ICU was associated with lower risk of being seropositive compared to those working in the emergency department/acute medicine.^4^ Our finding that nurses, NPs, and PAs had higher seroprevalence than physicians, although not statistically significant is empirically consistent with reports from some^8^ but not all prior studies.^4,21^ The timing of our study, which took place November 2020 – January 2021, is unique in comparison to the aforementioned studies which were generally conducted in the spring and summer of 2020 when community infection rates were much lower than was observed in our current study.

The high seroprevalence in January (44%) is likely driven by vaccination which started in mid-December, highlighting the early success of COVID-19 vaccination deployment among HCWs. Furthermore, as the time between dose 1 vaccination and serology testing increased, the seroprevalence increased as expected and in a manner consistent with reports from COVID-19 vaccine trials,^22^ suggesting that most individuals will be IgG positive by ∼14 days after the first vaccine dose. How this translates into immunity cannot be addressed in this study.

Our study included the following limitations: The results of this study are not generalizable to the larger population as we only enrolled a convenience sample of healthcare workers. As there is still limited information on the duration and variability of SARS-CoV-2 antibody response to infection, it is likely that our seroprevalence estimates are underestimates; specifically, we might have missed infections that occurred early in the pandemic for which antibody response waned or infections that occurred shortly before serology testing. Additionally, there are several potential sources of bias including information bias on reporting of previous infection with SARS-CoV-2 and recall bias related to vaccination dates, albeit this potential is minimized among healthcare professionals. Finally, the imperfect test sensitivity may bias the seroprevalence estimates, although we adjusted for testing error and found little impact on our estimates. It is also possible that information on setting and role, which was collected in April 2020 – June 2020, changed before participation in our seroprevalence studies conducted in November 2020 – January 2021.

Among a population of HCWs in the Minneapolis/St. Paul, MN metropolitan area, approximately one-in-six were infected with SARS-CoV-2 based on seropositivity. Seroprevalence ∼doubled during a three-month period between November 2020 and January 2021. Among unvaccinated individuals, physicians were at an empirically lower risk of being seropositive/previously infected. HCWs in ICU, inpatient, EMS, or emergency department settings experienced higher risk for being seropositive/previously infected. Given the high transmission rates and increasing hospitalization rates broadly occurring the U.S. at this time, future research that can inform optimal safety protocols for healthcare workers with direct patient contact will be important.

## Data Availability

De-identified data can be made available upon request to Dr. Demmer.

## ACKNOWLEDGEMENTS

*Financial support:* This study was supported by funding from the University of Minnesota Office of the Vice President for Research, by the Minnesota Population Center (funded by the Eunice Kennedy Shriver National Institute of Child Health and Human Development Population Research Infrastructure Program P2C HD041023), and by the The University of Minnesota’s NIH Clinical and Translational Science Award: UL1TR002494. Dr. Ulrich was supported by NIH grant T32AI05543315. We are also profoundly grateful for the study participants who have donated valuable time to advance our understanding about SARS-CoV-2 prevalence in healthcare workers.

